# Testosterone and socioeconomic position: Mendelian Randomization in 306,248 men and women participants of UK Biobank

**DOI:** 10.1101/2020.11.06.20226944

**Authors:** Sean Harrison, Neil M Davies, Laura D Howe, Amanda Hughes

**Affiliations:** MRC Integrative Epidemiology Unit, University of Bristol, Bristol, UK; Population Health Sciences, Bristol Medical School, University of Bristol, Bristol, UK; K.G. Jebsen Center for Genetic Epidemiology, Department of Public Health and Nursing, Norwegian University of Science and Technology, Norway

**Keywords:** testosterone, income, earnings, socioeconomic position, socioeconomic status, Mendelian randomization, UK Biobank

## Abstract

Men with more advantaged socioeconomic position (SEP) and better health have been observed to have higher levels of testosterone. It is unclear whether these associations arise because testosterone has a causal impact on SEP and health. In 306,248 participants of UK Biobank, we performed sex-stratified genome-wide association analysis to identify genetic variants associated with testosterone. Using the identified variants, we performed Mendelian randomization analysis of the influence of testosterone on socioeconomic position, including income, employment status, area-level deprivation, and educational qualifications; on health, including self-rated health and BMI, and on risk-taking behaviour. We found little evidence that testosterone affected socioeconomic position, health, or risk-taking. Our results therefore suggest it is unlikely that testosterone meaningfully affects these outcomes in men or women. Differences between Mendelian randomization and multivariable-adjusted estimates suggest previously reported associations with socioeconomic position and health may be due to residual confounding or reverse causation.

## Introduction

Testosterone has long been of interest in the study of human behaviour(1,2). For human and non-human primates, testosterone is thought to play a role in advancing and maintaining status compared to competitors(3,4). In experimental settings, it has been shown to promote either aggressive or prosocial behaviour depending on the context(5), and to influence economic decision-making, in particular financial risk taking(6–8). Outside the laboratory, there are reasons to believe that these same processes could over longer timescales impact people’s social and economic circumstances. Positive associations with circulating testosterone have for example been reported for self-employment, a financially ‘riskier’ strategy than standard employment(9,10), and likelihood of employment transitions(11). Work in male occupational samples points to a positive relationship of testosterone with aspects of socioeconomic position: among male executives, circulating testosterone has been linked with number of subordinates(12), and among male financial traders, with daily profits(13). Research has suggested that the in utero exposure to testosterone increases adult earnings for men(14), but a possible detrimental association for women(15)(16). However, correlates of testosterone exposure in utero and in adulthood may be very different, and observational studies of adults where testosterone and outcomes are measured together tell us little about causality. Adult testosterone may causally affect earnings, employment status, or other aspects a person’s socioeconomic position (SEP). At the same time, there are reasons to believe socioeconomic position may influence testosterone levels (i.e. reverse causation). Chronic stress can lower testosterone levels by affecting both production and secretion(17), suggesting psychosocial stress associated with socioeconomic adversity could influence testosterone alongside other aspects of health(18). SEP could also influence testosterone via obesity, which in men lowers circulating testosterone(19,20), and associates in high-income countries with disadvantage. Because smoking is more common in less advantaged groups but may raise testosterone, smoking could reduce rather than contribute to social differences in testosterone(21,22). Socioeconomic influences on testosterone could also be mediated by self-perception of social status, a dimension of SEP whose influences on health may be distinct from income(23). In competitive situations – both experimental settings and sports matches(24) - testosterone has been found to change depending on outcome, rising in the winner compared to the loser(2,25,26). Lastly, associations of testosterone with socioeconomic position could be explained by the relationship of testosterone with other aspects of health. For men, observational studies looking at naturally-occurring variation in testosterone have shown that higher testosterone is linked with better health, including lower risk of cardiovascular outcomes(27) and of diabetes(28,29), but also increased risk of prostate cancer(30). In women higher testosterone has been associated with poorer metabolic health, including cardiovascular outcomes(31), high blood pressure(32), and visceral adiposity(33). Higher circulating levels of testosterone and other androgens is also a key diagnostic feature of polycystic ovary syndrome(34). It is unclear how much these associations reflect impact of testosterone on health as opposed to impact of health on testosterone. Recent genetic evidence for an impact on cardiovascular health and associated risk factors(35)(36) does, however, indicate that testosterone is likely to have some causal impacts on health. This includes blood pressure, lipids(36), and risk of type 2 diabetes(35). One recent MR study suggested a beneficial influence on some aspects of health (bone mineral density, body fat) but a detrimental impact on others (high-density lipoprotein cholesterol, hypertension and prostate cancer risk)(37). Since health is known to influence diverse aspects of socioeconomic position(38), testosterone may therefore also impact SEP via health.

This evidence is however largely circumstantial, and despite plausible mechanisms, the relationship of testosterone and SEP is poorly understood. One issue is that most work has been experimental or involved specific occupational samples. Understanding associations more widely requires investigation in general population samples, since biological correlates of success for financial traders and sports players may not extend to the rest of the population. Secondly, both experimental and observational work has to date focused almost exclusively on men, and consequently little is known about how testosterone relates to socioeconomic circumstances in women. Thirdly, few studies have used causal inference methods to infer the effects of testosterone. Observational studies which do not are likely biased by confounding and reverse causation. This occurs even when a list of known confounders is included as covariates, since there may be substantial error in the measurement of those covariates, leading to ‘residual’ confounding. A UK study from 2015 found social differences in men’s circulating testosterone at age 60-64, with lower testosterone for men with lower income and fewer educational qualifications(39), but could not determine if this relationship was causal. A 2018 study in British men(40) applied Mendelian randomization, a genetic causal inference approach, to investigate the influence of testosterone on socioeconomic position. It found suggestive evidence of a positive influence on earnings and probability of being in work, but the sample size (N=3663) was insufficient to draw meaningful conclusions.

We investigate the impact of circulating testosterone on social and economic outcomes in 306,248 men and women from the UK Biobank. We investigated whether the associations of SEP and testosterone were likely to be causal using Mendelian randomization. Mendelian randomization uses genetic variants associated with an exposure of interest (usually single nucleotide polymorphisms, or SNPs) as instrumental variables for the exposure, in this case testosterone. Since SNPs are assigned at conception, associations with SNPs cannot be due to reverse causation or classical confounding of the exposure and the outcome (41). Multiple SNPs associated with an exposure can be combined into a polygenic score (PGS) representing overall genetic predisposition for the exposure. SNPs for the exposure are usually taken from an existing genome-wide association study (GWAS) based on a separate population to that in which the outcome is measured, because using the same population can result in bias (42). However, existing GWAS of testosterone not involving UK Biobank were based on small samples(43,44) which identified few SNPs. We therefore conducted a GWAS of testosterone in UK Biobank. We used a split-sample approach to delineate two independent samples (detailed in the methods section) thus avoiding bias caused by sample overlap(42). We examine the impact of testosterone on the following outcomes: employment status, household income, having a university degree, employment status, having a skilled job, area-level deprivation, home ownership, and partnership/cohabitation status. We also examine associations with factors proposed as mediators of testosterone-SEP associations: overall health, adiposity (BMI), and risk-taking behaviour. Building on previous work restricted to men, we examine all relationships in parallel for men and women separately.

## Methods

### Population

UK Biobank is a population-based health research resource consisting of over 500,000 people recruited between 2006 and 2010 from 22 centres across the UK(45). Participants provided socioeconomic information and anthropometric measures via questionnaires and interviews at recruitment, and biomarkers were measured using blood tests. The study design, participants and quality control methods have been described in detail previously(46–48). UK Biobank received ethics approval from the Research Ethics Committee (REC reference for UK Biobank is 11/NW/0382).

We included participants of white British ancestry with at least one measured blood level of: testosterone, sex hormone binding globulin (SHBG) or albumin. We excluded participants who reported at baseline that they were taking any drug that would influence their levels of testosterone or oestrogen, including hormone replacement therapy and the contraceptive pill; a full list of medications resulting in exclusion is available in **Supplementary Table 2**. We included only unrelated participants in all analyses. Full details of inclusion criteria and genotyping are in **Supplementary Information 1**. After exclusions, 306,248 unrelated participants remained in the dataset.

### Exposures

Testosterone circulates in several forms: unbound or ‘free’, loosely bound to albumin, or tightly bound to sex hormone binding globulin (SHGB). Bioavailable testosterone refers to the first two categories, which are available for biological processes and hence more relevant to purported causal effects. The primary exposure in this analysis is bioavailable testosterone, but in additional analyses we examined relationships with total and free testosterone. In UK Biobank, testosterone and SHBG were measured by one step competitive analysis on a Beckman Coulter Unicel Dxl 800 in nmol/litre, and albumin was measured by BCG analysis on a Beckman Coulter AU5800 in g/litre. There were 23 participants with the minimum detectable limit of 0.35 nmol/litre of testosterone; we coded these participants as having 0.35 nmol/litre of testosterone. All participants had over the minimum detectable levels of albumin and SHBG.

We estimated free testosterone using the method detailed in Ho et al. (2006) (49), based on the work of Vermeulen et al. (1999): (50).

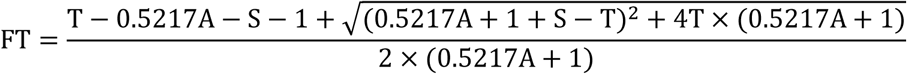

where FT is free testosterone, T is total testosterone (nmol/litre), S is SHBG (nmol/litre) and A is albumin (g/litre). Similarly, we estimated bioavailable testosterone as:

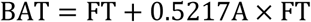

where BAT is bioavailable testosterone.

### Covariates

Age, sex and UK Biobank recruitment centre were reported at the baseline assessment, and 40 genetic principal components (used to control for population stratification (51)) were derived by UK Biobank.

We classified participants as having had an oophorectomy if they answered yes to the question “Have you had BOTH ovaries removed?”, and coded participants as having had menopause if they answered yes to “Have you had your menopause (periods stopped)?” or they had an oophorectomy. We coded time of blood collection as the number of hours from midnight to the time of first blood collection.

### Outcomes

**Box 2** describes outcomes, and **Supplementary Information 2** gives further information on their measurement. For household income and employment-related outcomes, we restricted the analysis to participants under the stage pension age at the time of recruitment (60 years for women and 65 years for men). Since these outcomes are closely linked with current labour market involvement, associations with testosterone would be expected to weaken considerably following retirement age. For all other outcomes, the full age range of UK Biobank was used.

#### Box 2

*List of main outcomes*

***Continuous outcomes***

- Average household income before tax, with each category assigned the mid-point of the range (and open-ended categories a nominal value) to allow for continuous analysis*:
  - <£18,000 = £15,000
  - £18,000 to £30,999 = £24,500
  - £31,000 to £51,999 = £41,500
  - £52,000 to £100,000 = £76,000
  - >£100,000 = £150,000
- Deprivation, measured using the Townsend Deprivation Index (TDI) of current address*
- Lifetime smoking behaviour, an index capturing initiation, heaviness and duration (52)
- Body mass index (BMI, kg/m^2^)

*We dichotomised income and deprivation as additional analyses: ≥£52,000 versus <£52,000 for income, most deprived third of TDI versus two least deprived thirds for deprivation

***Binary outcomes***

- Current employment status, coded as four separate outcomes:
  - Looking after home/family (homemaker) versus employed
  - Out of labour force (due to sickness/disability) versus employed
  - Retired versus employed
  - Unemployed versus employed
- Job class, coded as skilled versus unskilled (53)
- Degree status, coded as degree-level education versus lower
- Owner-occupied accommodation versus renting
- Whether in a cohabiting partnership
- Self-reported risk-taker versus not*
- Self-reported good or excellent overall health versus poor or fair overall health
- Current smoking status

Main analyses for household income and employment-related outcomes were restricted to participants under the state pension at recruitment (60 years for women, 65 years for men).

*Participants were asked: ‘Would you describe yourself as someone who takes risks?’ and could answer yes, no, do not know, or prefer not to say

#### Main Analysis

We analysed men, pre-menopausal women and post-menopausal women separately. Testosterone levels and genetic determinants differ substantially between men and women, and testosterone levels also differ between women who have and have not had an oophorectomy. We classified women who had reported having an oophorectomy as post-menopausal, and we excluded women for whom menopausal status was unknown from the main analyses.

As previous genome-wide association studies (GWAS) for bioavailable testosterone have been underpowered, we conducted new GWAS to identify genetic variants associated with bioavailable testosterone separately for men, pre-menopausal women, and post-menopausal women, retaining related participants. We used a split-sample approach, randomly splitting each of the three groups (men, pre-menopausal women, post-menopausal women) into two halves, or ‘splits’, and conducting a GWAS in each split using the MRC IEU UK Biobank GWAS pipeline(54). For men, pre-menopausal and post-menopausal women, we then performed MR analyses separately in each of the two splits, using the SNPs identified in the GWAS of the other. Conducting a split-sample analysis avoids bias from using overlapping GWAS and analytic samples(42), while maintaining statistical power. In all GWAS, we used BOLT-LMM (55)(which accounts for relatedness between participants and population structure), with age and 40 principal components as covariates. For post-menopausal women, we also included having had an oophorectomy as a covariate.

For men, pre-menopausal women and post-menopausal women, we clumped the GWAS significant SNPs (P < 5 x 10^−8^) from each split’s GWAS with a clumping window of 10,000 kb and an R^2^ threshold of 0.001. We used the clumped SNPs to create a polygenic score (PGS) for bioavailable testosterone in unrelated participants in the other split. We calculated the PGS as the sum of each individual’s testosterone-increasing alleles weighted by the regression coefficient from the GWAS.

Within each split, we used Mendelian randomization to estimate the causal effect of bioavailable testosterone on all outcomes, using the PGS as an instrumental variable, with age at baseline assessment (linear, squared and cubed), time to blood collection (linear, squared and cubed), UK Biobank recruitment centre, and 40 genetic principal components as covariates. We used the ivreg2 package in Stata (version 15.1) with robust standard errors, and tested for weak instrument bias (using F statistics) to assess whether the PGS were sufficiently predictive of testosterone (56).

Mendelian randomization analyses estimate mean and risk differences for continuous and binary outcomes respectively using additive structural mean models(57–59). Mean differences are interpreted as the average change in a participant’s outcome (e.g. household income) per unit increase in the exposure. Risk differences are interpreted as the absolute percentage point change in the proportion of participants with the outcome (e.g. unemployment) per unit increase in the exposure (as in a linear probability model).

For men, pre-menopausal women and post-menopausal women, we used fixed-effect meta-analysis to combine the results from the two splits, giving a final estimate of the effect of bioavailable testosterone on each outcome. We express all results per standard deviation (SD) increase in bioavailable testosterone, estimating the SD of bioavailable testosterone within men, pre-menopausal and post-menopausal women separately.

To compare the Mendelian randomization results with associations from multivariable adjusted analysis, within each split we estimated multivariable adjusted associations between bioavailable testosterone and all outcomes using linear regression, with age at baseline assessment (linear, squared and cubed), UK Biobank recruitment centre, time to blood collection (linear, squared and cubed), and 40 genetic principal components as covariates, i.e. observational analyses without genetic instruments. We used linear probability models for binary outcomes rather than logistic regression models, to be comparable with the Mendelian randomization analyses. We also performed used Hausman tests (60) within each split to test whether estimates from Mendelian randomization and multivariable adjusted analyses differed. As Hausman tests are only possible to conduct within each split, we used Fisher tests(61) to test whether estimates from the Mendelian randomization and multivariable adjusted analyses differed after meta-analysis. All statistical tests were two-tailed.

#### Sensitivity Analyses

Mendelian randomization analysis is based on the assumption that SNPs, and therefore PGSs, are not pleiotropic, i.e. that they do not affect the outcome except through the exposure. We conducted sensitivity Mendelian randomization analyses to test this assumption, including inverse-variance weighted (IVW), MR-Egger (which provides a test of certain forms of pleiotropy), weighted median and weighted mode analyses(62–64). We also measured Cochran’s Q statistic from the IVW analyses (a measure of heterogeneity in the estimated effects on outcome using individual SNPs), an indicator of pleiotropy (65) or problems with modelling assumptions(66). We combined the results of each analysis from each split using fixed effect meta-analysis, as in the main analysis.

We determined from these analyses a) if results of the sensitivity analysis were consistent with those from the main Mendelian randomization analysis, indicating robustness of the main results, and b) whether there was evidence of pleiotropy, from both the Egger regression constant term and Cochran’s Q statistic. We also inspected plots showing the results of the sensitivity Mendelian randomization analyses, which would indicate possible bias in the results of the main analysis.

Further information for the sensitivity analyses and results are in **Supplementary Information 3**.**1**.

#### Secondary Analyses

We repeated the GWAS, and all subsequent analyses, for total testosterone, free testosterone, albumin and SHBG. We also repeated the GWAS and all analyses for all women regardless of menopausal status (adjusting the GWAS and analyses for menopause [categorical: yes, no, don’t know] and oophorectomy), and additionally for post-menopausal women but without adjusting for oophorectomy.

As additional outcomes, we included participants above retirement age in household income and all employment variables, as well as equivalised household income, defined as household income divided by the number of people living in the same household, setting the maximum number in the household at 12.

Further information for the secondary analyses and results are in **Supplementary Information 3**.**2**.

#### Patient and Public Involvement

This study was conducted using UK Biobank. Details of patient and public involvement in the UK Biobank are available online (www.ukbiobank.ac.uk/about-biobank-uk/ and https://www.ukbiobank.ac.uk/wp-content/uploads/2011/07/Summary-EGF-consultation.pdf?phpMyAdmin=trmKQlYdjjnQIgJ%2CfAzikMhEnx6). No patients were specifically involved in setting the research question or the outcome measures, nor were they involved in developing plans for recruitment, design, or implementation of this study. No patients were asked to advise on interpretation or writing up of results. There are no specific plans to disseminate the results of the research to study participants, but the UK Biobank disseminates key findings from projects on its website.

#### Data and Code Availability

The empirical dataset will be archived with UK Biobank and made available to individuals who obtain the necessary permissions from the study’s data access committees. The code used to clean and analyse the data is available in supplementary materials, and here: https://github.com/sean-harrison-bristol/Testosterone_and_sep

## Results

Summary demographics are presented in **Table 1**. Briefly, we analysed 148,248 men, 36,203 pre-menopausal women and 104,632 post-menopausal women. The mean bioavailable testosterone level was 5.21 nmol/litre in men (standard deviation [SD]: 1.54 nmol/litre), 0.37 nmol/litre in pre-menopausal women (SD: 0.25 nmol/litre), and 0.36 nmol/litre in post-menopausal women (SD: 0.28 nmol/litre). The PGS for bioavailable testosterone in each split had R^2^ values of 3.3% (from 41 SNPs) and 3.5% (from 40 SNPs) for men, 0.7% (from 6 SNPs) and 0.7% (from 7 SNPs) for pre-menopausal women, and 0.04% (from 76 SNPs) and 0.4% (from 5 SNPs) for post-menopausal women; **Supplementary Table 3** details the PGS for all exposures, and **Supplementary Table 4** details all the GWAS-significant SNPs from the GWAS.

**Table 1:**
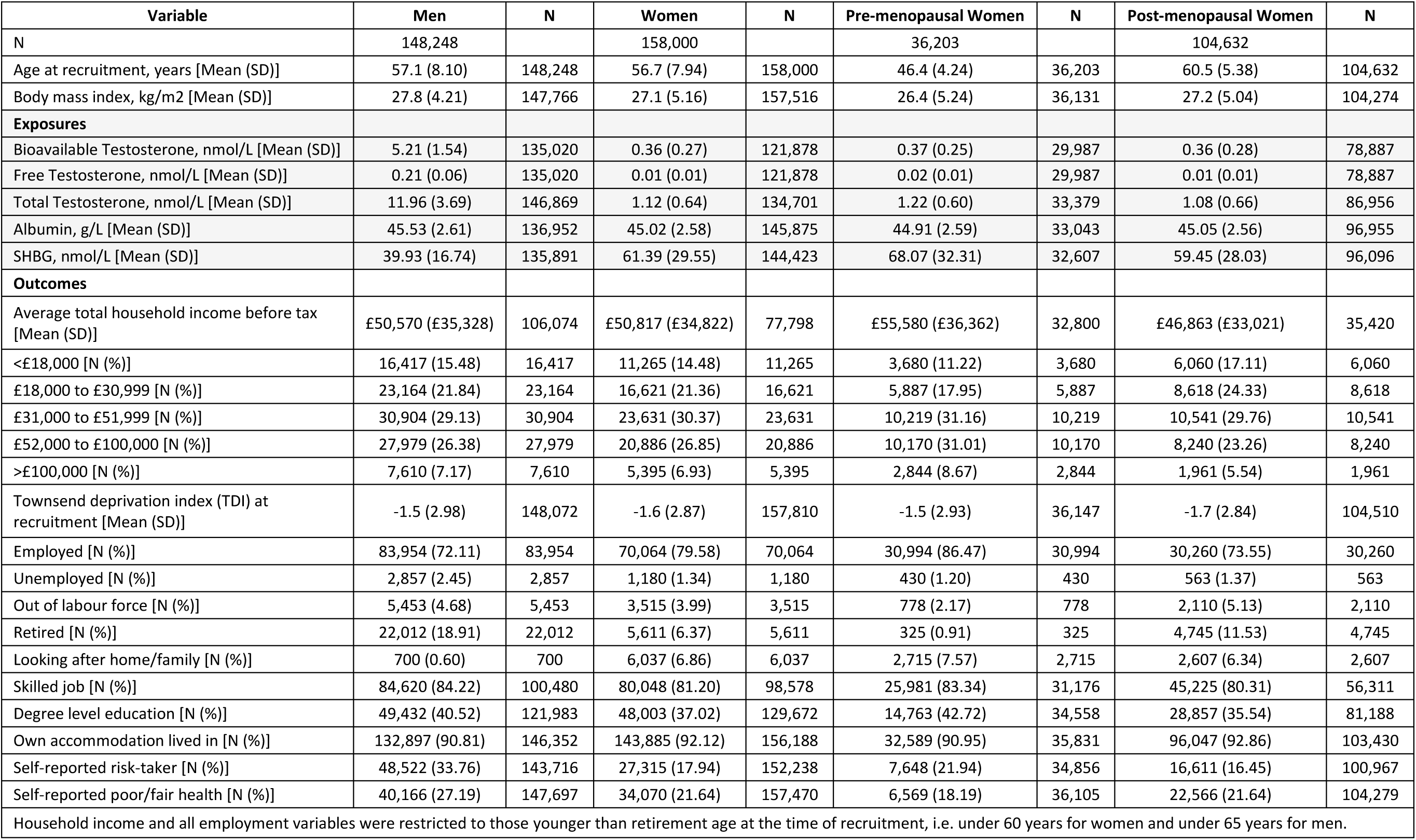
Summary demographics.

**Supplementary Table 1** shows results from the Mendelian randomization and multivariable adjusted analyses for all outcomes, while **Supplementary Table 5** shows results from all main and secondary analyses. Forest plots showing the Mendelian randomization and multivariable adjusted analysis results for all binary outcomes are shown in **Figures 1**-**3**. Mendelian randomization and multivariable adjusted analysis results for continuous outcomes are shown in **Figures 4 and 5**. Forest plots for other exposures are available in **Supplementary Materials**.

**Figure 1:**
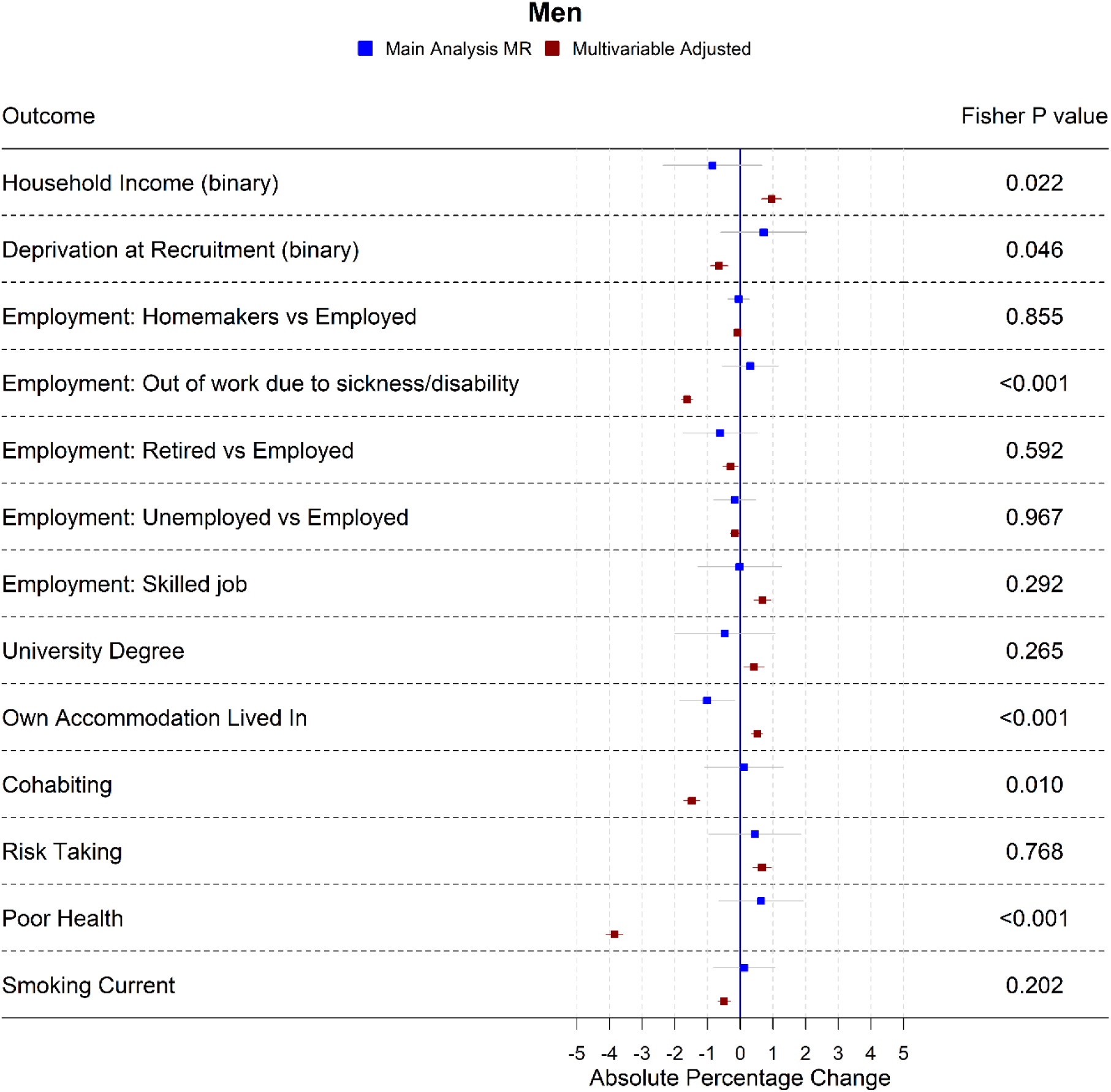
Forest plot showing the effect of bioavailable testosterone on all binary outcomes for men. Note: employment outcomes were restricted to men younger than 65 years at the time of recruitment

### Bioavailable testosterone in men

In multivariable-adjusted models, higher bioavailable testosterone in men was associated with more advantaged SEP (**Figure 1, Figure 4, Supplementary Table 1**). For instance, a one-SD increase in bioavailable testosterone corresponded to £851 (95%CI: £628 to £1075) greater annual household income, greater likelihood of having a university degree (absolute percentage change [APC] 0.42%, 95%CI: 0.11% to 0.73%), and greater likelihood of holding a skilled job (APC = 0.68%, 95%CI: 0.41% to 0.94%). Bioavailable testosterone was also positively associated with health (**Figure 1, Figure 5, Supplementary Table 1**). For instance, a one-SD increase in bioavailable testosterone corresponded to a 0.50 kg/m^2^ lower BMI (0.47 to 0.52) and lower probability of reporting poor health (APC =-3.85% (−4.11% to -3.60%). It was negatively associated with partnership (APC =-1.48%, CI: -1.72% to -1.24%), positively associated with self-assessed risk taking 0.66% (APC =-0.39% to 0.94%), and negatively associated with smoking, for example a corresponding to a decrease in the lifetime smoking index of 0.06 (−0.06 to -0.05).

In Mendelian randomization analyses (**Figures 1, 4 and 5, Supplementary Table 1**), effect sizes were smaller and confidence intervals larger. There was little evidence for causal effects of bioavailable testosterone on any outcome, with all 95% confidence intervals crossing the null. For instance, a one-SD increase in men’s bioavailable testosterone corresponded to difference in annual household income of -£31 (95%CI: -£1147 to £1086), an increase in BMI of 0.06 kg/m^2^ (95%CI: -0.06 to 0.19) and a difference in the lifetime smoking index of 0.02 (95%CI:-0.01 to 0.05). The absolute percentage change (APC) for having a university degree was -0.47% (95%CI: -2.00% to 1.06%), for likelihood of holding a skilled job, -0.02% (95%CI: -1.30% to 1.25%), for reporting poor health 0.64% (−0.66% to 1.94%) and for partnership 0.11%(−1.08% to 1.31%). Although confidence intervals were wide, we were able exclude as very unlikely effect sizes beyond the upper and lower confidence limits of these estimates: for example, an impact of a one-SD increase in bioavailable testosterone on annual household income greater than £1086, or a decrease in BMI greater than 0.06kg/m^2^. Fisher tests (**Figures 1, 4 and 5, Supplementary Table 1**) gave evidence that for several SEP and health-related outcomes, differences between the multivariable-adjusted and Mendelian randomization estimates were substantively different. This was the case for **the chance of being out of work due to sickness or disability** (p=1.02×10^−5^), **the chance of owning accommodation** (p=4.28×10^−4^), **deprivation at recruitment** (p = 0.002), as well as **the chance of self-reporting poor health** (p = 2.91 x 10^−11^), **BMI** (p= 1.56 x 10^−18^) and **lifetime smoking** (p = 4.12×10^−6^).

The PGS for bioavailable testosterone was not correlated with either the PGS for albumin or the PGS for SHBG (see **Supplementary Table 7**). There was little consistent evidence in both splits of heterogeneity between SNPs in the sensitivity Mendelian randomization analyses, and little evidence of pleiotropy from MR-Egger regression (**Supplementary Table 6)**. There was also a low risk of weak instrument bias for men; the F-statistics in all analyses were above 1000.

### Bioavailable testosterone in pre-menopausal women

In multivariable-adjusted models, bioavailable testosterone among pre-menopausal women was generally associated with less advantaged socioeconomic position (**Figures 2 and 4, Supplementary Table 1**). For instance, a one-S.D. increase in bioavailable testosterone corresponded to lower annual household income (−£1752, 95% CI: -£2171 to -£1332), lower probability of having a degree (APC = -1.99%, 95%CI: -2.55% to -1.42%), and lower probability of owning own accommodation (APC =-1.12%, 95%CI: -1.44% to -0.79%). It corresponded to a 1.60kg/m^2^ (95%CI: 1.54 to 1.66) higher BMI and was positively associated with smoking, for instance with an increase in the lifetime smoking index of 0.04(95%CI: 0.04 to 0.05) (**Figure 5, Supplementary Table 1**). As for men, bioavailable testosterone was negatively associated with partnership (APC per S.D. increase = -1.53%, 95%CI: -2.03% to -1.02%), and positively associated with self-assessed risk-taking (APC = 0.53%, 95%CI: 0.05% to 1.01%).

**Figure 2:**
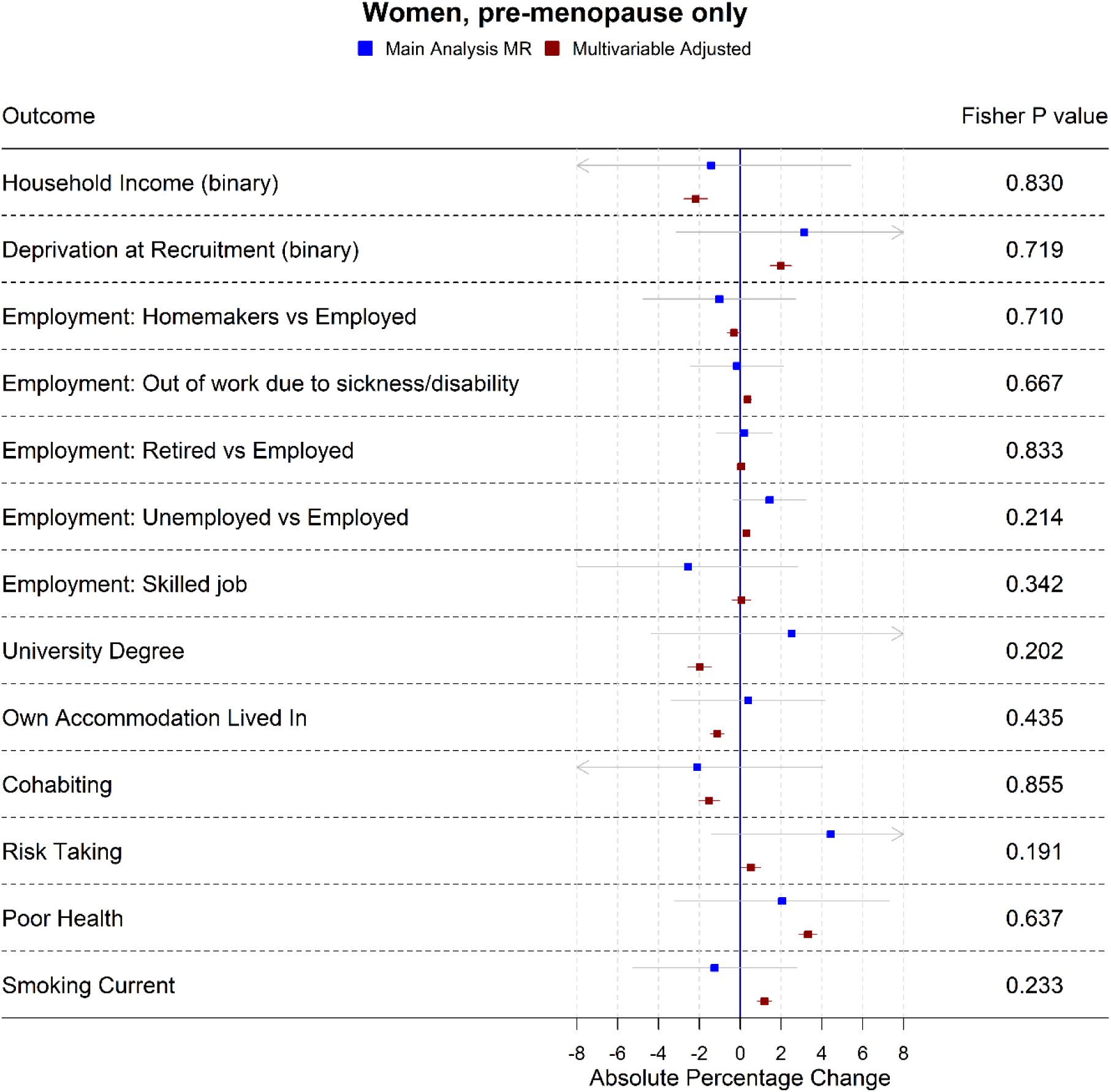
Forest plot showing the effect of bioavailable testosterone on all binary outcomes for pre-menopausal women. Note: employment outcomes were restricted to women younger than 60 years at the time of recruitment

In Mendelian randomization analyses (**Figures 2, 4 and 5, Supplementary Table 1**), there was little evidence for any causal effects of bioavailable testosterone for pre-menopausal women. Differences per one-S.D. increase in bioavailable testosterone were: -£1583 for annual household income (95% CI: -£6598 to -£3432), 0.01kg/m^2^ for BMI (95%CI: -0.72 kg/m^2^ to 0.74 kg/m^2^) and -0.08 (95%CI:-0.19 to 0.03) for the lifetime smoking index. For probability of having a degree, owning one’s own accommodation, and partnership, APCs were 2.51% (95%CI: -4.37% to 9.40%), 0.39%(95%CI: -3.38% to 4.16%), and -2.10%(95%CI: -8.24% to 4.04%). For self-assessed risk taking, it was 4.43% (95%CI: -1.40% to 10.26%). Confidence intervals were wider than for men, but a **Fisher test** (p= 2.04×10^−5^) **indicated that for BMI** the Mendelian randomization estimate (beta = 0.01 kg/m^2^, 95% CI: -0.72 kg/m^2^ to 0.74 kg/m^2^) differed from the multivariable-adjusted estimate. This was also the case for lifetime smoking (p=0.007) and current smoking (p=0.004). The PGS for bioavailable testosterone was among pre-menopausal women negatively correlated with the PGS for SHBG (r = -0.39), see **Supplementary Table 7**. F-statistics in all analyses were above 85, indicating that for pre-menopausal women there was a low risk of weak instrument bias.

### Bioavailable testosterone in post-menopausal women

In post-menopausal women, bioavailable testosterone was negatively associated in multivariable-adjusted models with several measures of SEP (**Figures 3 and 4, Supplementary Table 1**). A one-S.D. increase in bioavailable testosterone corresponded to £996 (£-£600 to £1391) lower annual household income, lower probability of owning own accommodation (APC = 0.33%, 95%CI: -0.51% to -0.15%) and higher TDI at recruitment 0.04 (0.02 to 0.06). It corresponded to a 1.19 kg/m^2^ (95%CI: 1.16 to 1.23) higher BMI, more lifetime smoking (beta = 0.02, 95%CI:0.01 to 0.03), and greater likelihood of reporting poor health (APC = 1.85%, 95%CI: 1.57% to 2.14%). Unlike for men and for pre-menopausal women, bioavailable testosterone was positively associated with cohabitation (APC = 0.33%, 95%CI: 0.01% to 0.64%), and there was little evidence of an association with risk-taking (APC = -0.26%, 95%CI: -0.52% to 0.00%).

**Figure 3:**
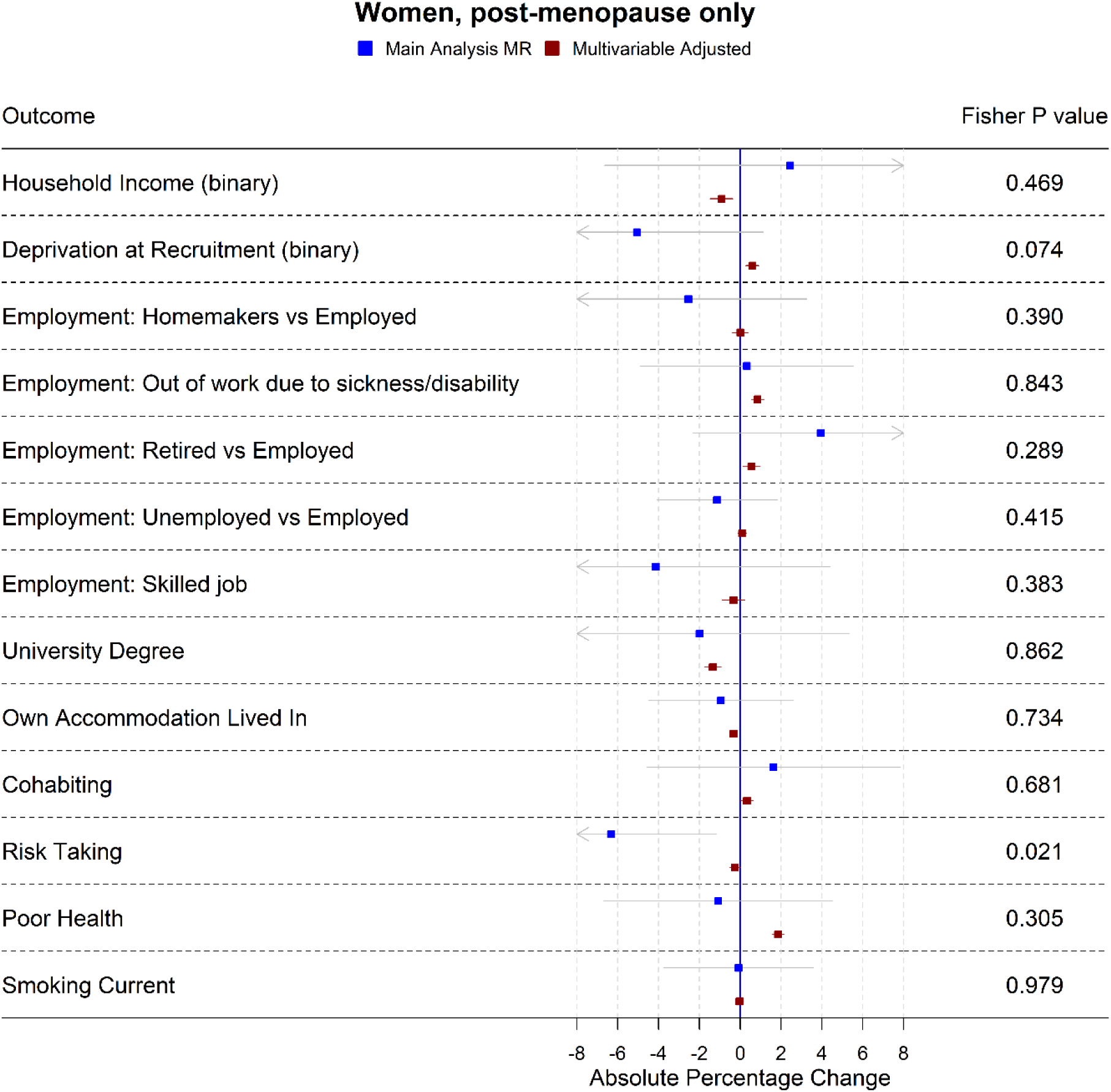
Forest plot showing the effect of bioavailable testosterone on all binary outcomes for post-menopausal women. Note: employment outcomes were restricted to women younger than 60 years at the time of recruitment

The Mendelian randomization estimates (**Figures 3-5, Supplementary Table 1**) provided little evidence of a causal effect on most outcomes. Differences per one-S.D. increase in bioavailable testosterone were for annual household income -£2063 (−£8346 to £4221), for BMI -0.43 kg/m^2^ (95%CI: -1.14 kg/m^2^ to 0.29 kg/m^2^), and for TDI at recruitment -0.32(95%CI:-0.69 to 0.05). For likelihood of owning one’s accommodation, of reporting poor health, and partnership, APCs were -0.94% (95%CI: -4.49% to 2.60%), -1.08% (−6.67% to 4.52%) and 1.63% (95%CI: -4.56% to 7.82%). For BMI, the Fisher test showed the MR and multivariable-adjusted estimates were different (p=9.19×10^−6^). A negative association for which the confidence interval did not cross the null was seen with the lifetime smoking index -0.15 (−0.28 to -0.02) and self-assessed risk-taking -6.33% (−11.49% to -1.17%). In both cases, Fisher tests (p=9.19×10^−6^, p=0.02) gave evidence for a substantive difference between these estimates and the multivariable-adjusted estimates. In post-menopausal women the PGS for bioavailable testosterone was negatively correlated with PGS for SHBG (r = -0.25), see **Supplementary Table 7**. F-statistics for most analyses were above 10: only some employment outcomes for post-menopausal women were not above 10. Employment outcomes for post-menopausal women may therefore have a higher chance of weak instrument bias.

**Figure 4:**
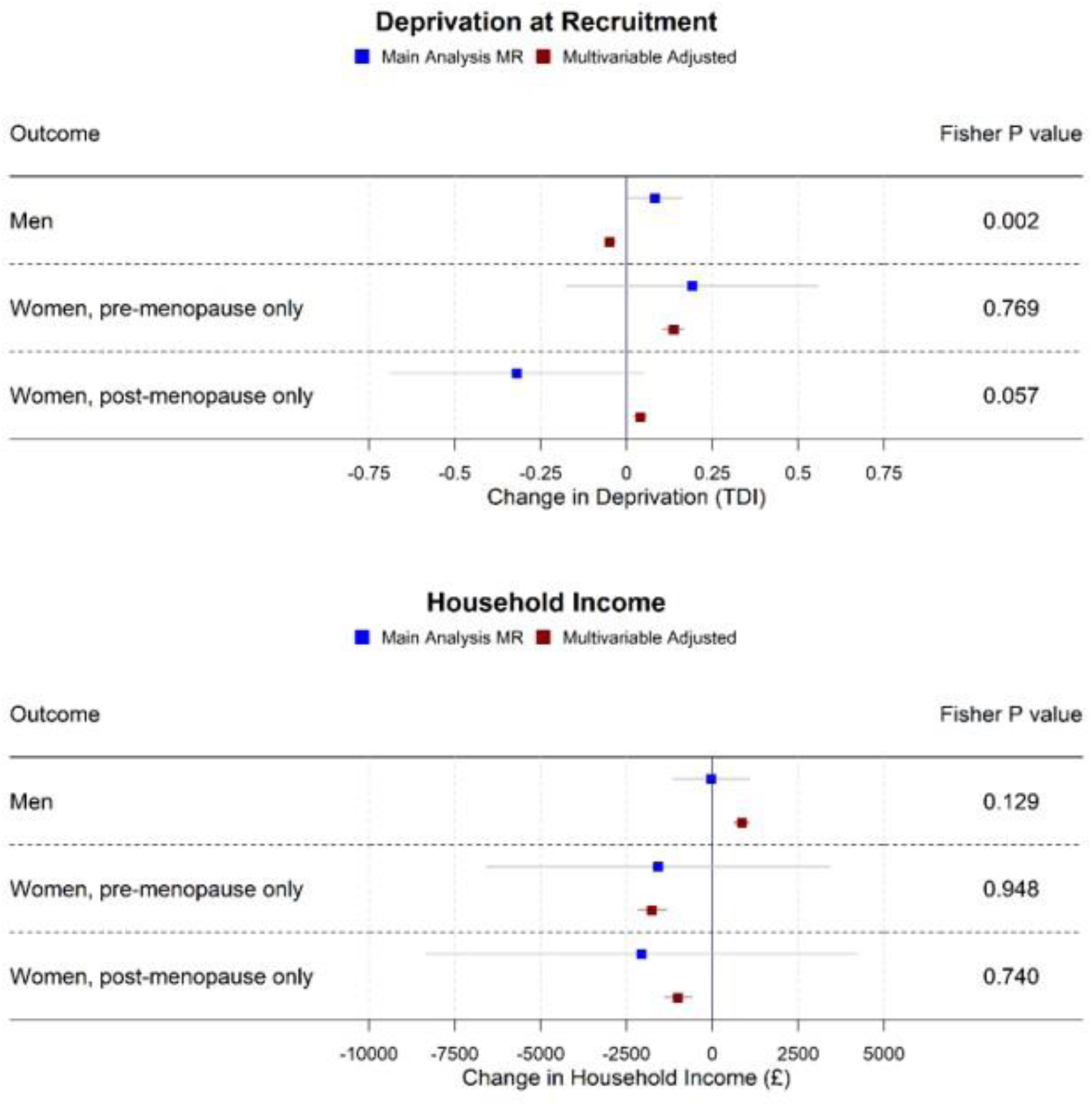
Forest plot showing the effect of bioavailable testosterone on continuous SEP outcomes for men, pre-menopausal women, and post-menopausal women.

**Figure 5:**
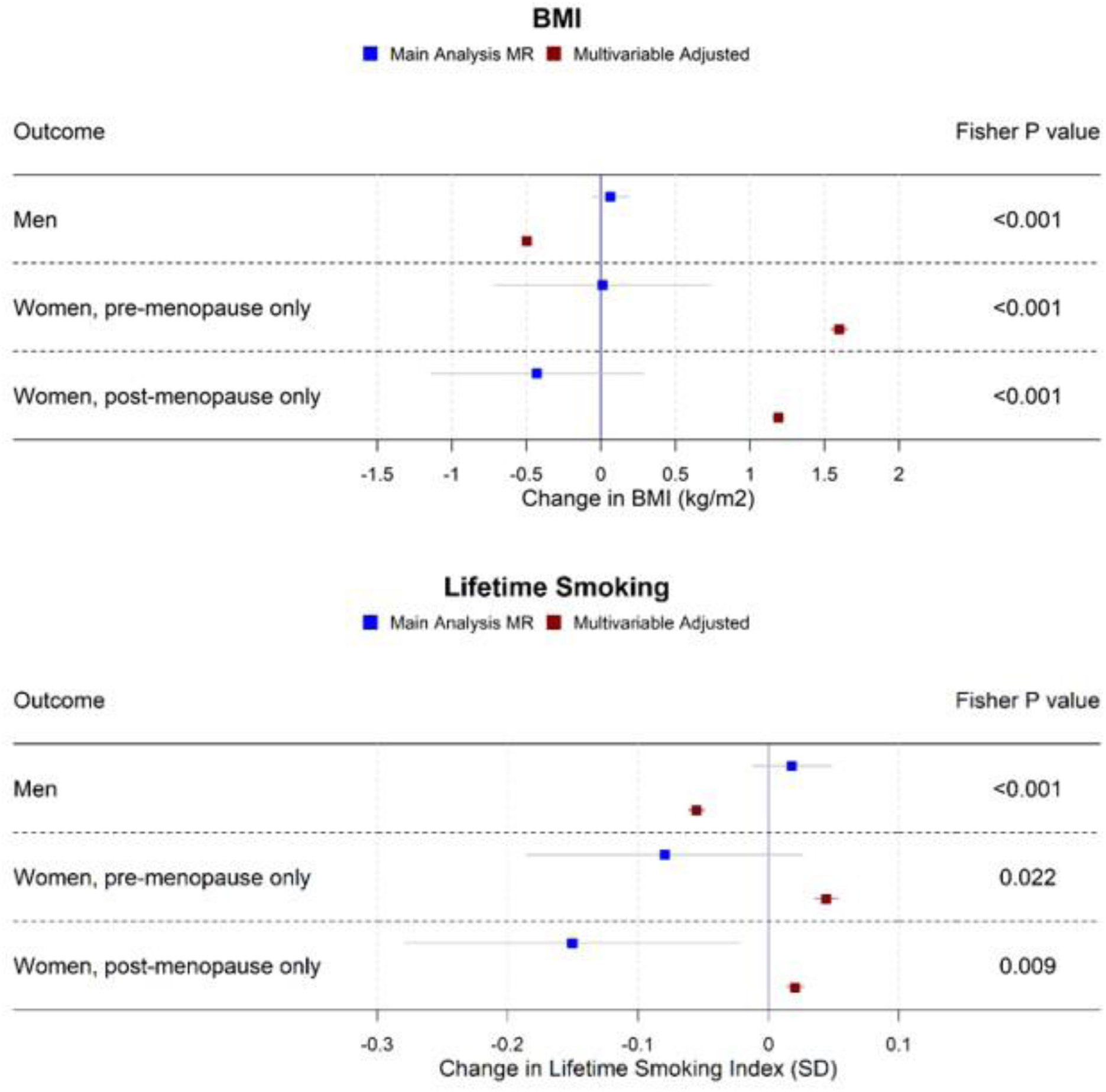
Forest plot showing the effect of bioavailable testosterone on continuous health outcomes for men, pre-menopausal women, and post-menopausal women.

## Discussion (no subheadings)

Consistent with existing observational work(39)(12)(14)(27)(31), multivariable-adjusted regression models found that bioavailable testosterone was associated with more advantaged socioeconomic position and better health among men, but less advantaged socioeconomic position and worse health for women. In contrast, the Mendelian randomization estimates provided little evidence that testosterone had causal effects on SEP or health. Fisher tests provided substantial evidence of heterogeneity between the multivariable adjusted and Mendelian randomization estimates. The Mendelian randomization estimates were precise enough to suggest that even small effects of testosterone on SEP or health are unlikely. Since Mendelian randomization analyses are not affected by classical confounding or reverse causation of the exposure-outcome relationship, this suggests that many previously reported associations of testosterone with socioeconomic outcomes, self-rated health and BMI are unlikely to be causal. Unlike our study, a recent Mendelian randomization analysis(35) supported in a causal impact of bioavailable testosterone on women’s BMI, although confidence intervals for the two studies’ results overlapped. This may reflect different analytic decisions – for instance, that study conducted a GWAS in the whole UKB population, with outcome information taken from a different study population entirely. Another(37) supported an impact on body fat in men. Since in that study the identification of genetic instruments and the Mendelian randomization were conducted in the same sample, without applying a split-sample approach, results may in part reflect bias due to sample overlap(42).

Risk-taking was in multivariable-adjusted regression models positively associated with testosterone for men and pre-menopausal women. The Mendelian randomization estimates provided little evidence of a positive relationship, but some evidence of a negative relationship for post-menopausal women. This runs contrary to evidence from experimental settings(6–8) supporting a positive association of testosterone with risk-taking behaviour. It could reflect lack of generalizability beyond experimental settings, or the limitations of measuring risk-taking with a self-reported and subjective measure.

A previous Mendelian randomization study by Hughes and Kumari(40) found suggestive support for an impact of testosterone on men’s earnings and men’s probability of being employed, but the sample size was small (N=3663) and estimates too imprecise to draw firm conclusions. UKB collected information on household income, but not individual earnings. It is possible that causal influence of testosterone on income relates to individual labour income specifically, although the current study found no evidence for an impact on men’s employment status either. Of note, in that study testosterone was instrumented with three SNPs, the only variants at the time known to associate with testosterone. The most predictive of these was not available in the current study (nor were any SNPs which could be used as proxies). Since the current study used more SNPs, we were able to apply methods to investigate pleiotropy. For this reason, also, the current study may be considered more robust.

### Strengths and Limitations

The main strengths of this analysis are that Mendelian randomization analyses are not affected by classical kinds of confounding or reverse causation which can affect other observational methods even when covariates are adjusted for(67). Given plausible mechanisms for an impact of socioeconomic position on testosterone, as well as the reverse, this is an important consideration. The UK Biobank is much larger than the studies previously used in this area – our analytic sample was approximately 84 times larger than that of the previous MR study on the topic(40). This allowed us to examine multiple health exposures and multiple socioeconomic and social outcomes with greater precision than previously possible. For some associations, there were marked differences between the Mendelian randomization and multivariable adjusted estimates, which suggests the multivariable adjusted estimates may suffer from reverse causation or residual confounding. Additionally, owing to the split-sample GWAS design, the SNPs contributing to the PGS were not biased by sample overlap(42), did not suffer from heterogeneity between the GWAS and analysis dataset, and all SNP reached genome-wide significance.

Risk-taking behaviour was based on a subjective self-reported measure and may not adequately capture behaviour relevant to testosterone. Recent evidence suggests that testosterone and cortisol jointly influence risk-taking(8), but measurements of cortisol were not available. Mendelian randomization rests on assumptions which are difficult to test(67). Robustness checks including IVW, MR-Egger, weighted median and weighted mode regression found little evidence of pleiotropy, but bias due to pleiotropy cannot be ruled out. The number of SNPs making up the PGS for bioavailable testosterone was low for pre-menopausal and post-menopausal women, limiting statistical power. The PGS for post-menopausal women included many more SNPs in split 1 (SNPs = 76) than in split 2 (SNPs = 5), despite explaining far less of the variance (R^2^ = 0.04% in split 1, R^2^ = 0.4% in split 2). This is likely due to chance, as almost all the SNPs included in split 1 have small minor allele frequencies. This meant that small numbers of post-menopausal women with atypical bioavailable testosterone values may have overly-influenced the GWAS in split 1, leading to the inclusion of more SNPs that were less predictive of bioavailable testosterone. The results of post-menopausal women may therefore be less robust than for men and pre-menopausal women. The PGS represents lifetime exposure to bioavailable testosterone, and effects at specific points in life cannot be explored with the methodology used in this paper. Another important potential source of bias in Mendelian randomization analyses is family-level genetic effects, for example the impact of parents’ genes on offspring via environmental pathways, known as dynastic effects or genetic nurture(68). Recent evidence suggests that these processes are especially relevant to Mendelian randomization with socioeconomic outcomes, for instance substantially distorting estimates of the causal impact of BMI on educational attainment(69). This may also occur for socioeconomic effects of testosterone. Methods robust to these biases exist but require data on large numbers of related individuals. These ‘within-family’ Mendelian randomization methods have been applied in UK Biobank using a stronger genetic instrument than was available here, but estimates were too imprecise to draw conclusions(70). For this research question, analysis would need to be further restricted to same-sex sibling pairs. We have therefore not used within-family analyses in this paper, as power would be extremely limited. However, it should be noted that these phenomena tend to inflate estimated causal effects rather than push them towards the null. Our MR results were consistent with the null and, for men, relatively precise. These null findings are therfore unlikely to be due to ancestary, dynastic effects, or assortative mating.

Participants of UK Biobank tend to be wealthier and healthier than the country as a whole, and this non-representativeness may bias estimates(71). An additional source of potential bias is geographic structure in the UK Biobank genotype data. Frequencies of genetic variants differ between ancestral populations and hence between parts of the UK, but so do environmental and cultural factors which influence traits of interest independently of genetics. This may cause confounding which cannot be accounted for by adjusting for principal components(72). However, recent evidence suggests that while geographic structure may be present after controlling for principal components in the PGS for exposures associated with educational attainment (e.g. BMI, smoking), there was little evidence for geographic structure in other PGS(73), implying bioavailable testosterone may be less subject to this bias. Additionally, a recent GWAS of income showed that only 8% of the inflation in the GWAS test statistics was due to residual stratification or confounding(74), indicating that population structure is unlikely to severely bias many of our results. As with family-level effects, while these effects could explain a false positive, they are unlikely to explain our negative results.

In conclusion, application of genetic causal inference methods in a large UK sample suggest previously reported associations of testosterone with socioeconomic outcomes, health, and risk-taking are unlikely to be causal.

## Data Availability

The empirical dataset will be archived with UK Biobank and made available to individuals who obtain the necessary permissions from the UK Biobank data access committees. The code used to clean and analyze the data is available in supplementary materials and on github via the link provided.

https://github.com/sean-harrison-bristol/Testosterone_and_sep

## Funding and Acknowledgements

This work was supported by the Health Foundation as part of a project entitled ‘social and economic consequences of health: causal inference methods and longitudinal, intergenerational data’, which is part of the Health Foundation’s Social and Economic Value of Health programme (Grant ID: 807293). The Health Foundation is an independent charity committed to bringing about better health and health care for people in the UK. LDH is funded by a Career Development Award from the UK Medical Research Council (MR/M020894/1). The Medical Research Council (MRC) and the University of Bristol support the MRC Integrative Epidemiology Unit [MC_UU_12013/1, MC_UU_12013/9, MC_UU_00011/1]. The Economics and Social Research Council (ESRC) support NMD via a Future Research Leaders grant [ES/N000757/1] and a Norwegian Research Council Grant number 295989. This publication is the work of the authors, who serve as the guarantors for the contents of this paper.

## Author Contributions

LDH and NMD obtained funding for this study. SH cleaned and analysed the data. SH and AH wrote the first draft. All authors contributed to study design, interpreted the results and revised the manuscript.

## Transparency Statement

Transparency statement: The lead author (the manuscript’s guarantor) affirms that this manuscript is an honest, accurate, and transparent account of the study being reported; that no important aspects of the study have been omitted; and that any discrepancies from the study as planned (and, if relevant, registered) have been explained.

