## Supplementary Information for "Testosterone and socioeconomic position: Mendelian Randomization in 306,248 men and women participants of UK Biobank"

### 1. Inclusion Criteria and Genotyping

#### Inclusion Criteria

We restricted analyses to individuals of white British ancestry, as defined by participants who self-reported as "White British" and who had very similar ancestral backgrounds according to the principal component analysis (n=409,703), as described by Bycroft (1). We excluded individuals with sex-mismatch (derived by comparing genetic sex and reported sex) or individuals with sex-chromosome aneuploidy from the analysis (n=814). We estimated kinship coefficients using the KING toolset (2) and identified 107,162 pairs of related individuals (1). We applied an in-house algorithm to this list and preferentially removed the individuals related to the greatest number of other individuals until no related pairs remain. This resulted in the exclusion of 79,448 individuals from the Mendelian randomization analyses (though related participants were included in the split-sample GWAS, as relatedness was accounted for). Additionally, two individuals were removed due to them relating to a very large number (>200) of individuals, and 135 individuals were excluded as they withdrew from the study.

Additionally, we excluded 339 participants who reported taking a testosterone altering drug, 5,108 participants who reported taking an oestrogen altering drug, 2,451 participants who reported taking the contraceptive pill, and 5,849 participants who reported taking hormone replacement therapy, a total of 13,105 participants, as some participants reported taking more than one testosterone or oestrogen altering drug. We also excluded 17,644 participants who had no measured levels of albumin, sex hormone binding globulin (SHBG) and testosterone.

After exclusions, 370,976 (related) participants remained for the split-sample GWAS, and 306,248 unrelated participants remained for the Mendelian randomization analyses.

#### Genotyping

The full data release contains the cohort of successfully genotyped samples (n=488,377). 49,979 individuals were genotyped using the UK BiLEVE array and 438,398 using the UK Biobank axiom array. Pre-imputation QC, phasing and imputation are described elsewhere (1). In brief, prior to phasing, multiallelic SNPs or those with MAF ≤1% were removed. Phasing of genotype data was performed using a modified version of the SHAPEIT2 algorithm (3). Genotype imputation to a reference set combining the UK10K haplotype and HRC reference panels (4) was performed using IMPUTE2 algorithms (5). The analyses presented here were restricted to autosomal variants within the HRC site list using a graded filtering with varying imputation quality for different allele frequency ranges. Therefore, rarer genetic variants are required to have a higher imputation INFO score (Info>0.3 for minor allele frequency (MAF) >3%; Info>0.6 for MAF 1-3%; Info>0.8 for MAF 0.5-1%; Info>0.9 for MAF 0.1- 0.5%) with MAF and Info scores having been recalculated on an in house derived 'European' subset.

Further information on the MRC-IEU quality control of UK Biobank genetic data is available online (6).

### 2. Outcome Definitions

#### Household Income and Deprivation

We recoded household income from categories to numerical values by taking the mid-point of the range, or a nominal value for the open-ended categories:

- <£18,000 = £15,000
- £18,000 to £30,999 = £24,500
- £31,000 to £51,999 = £41,500
- £52,000 to £100,000 = £76,000
- >£100,000 = £150,000

We created binary variables for household income and deprivation (as measured by Townsend Deprivation Index [TDI]), which we analysed along with the continuous measures. For household income, we compared those with a total household income above and below £52,000, i.e. upper two categories of household income versus bottom three categories. For deprivation, we split the participants into tertiles of TDI, and compared the most deprived tertile with the remaining two tertiles. These results are included in forest plots of results, but not included in **Supplementary Table 1.**

#### Employment

We coded job class as skilled versus unskilled as in Tyrrell (2016, (7)), where a skilled job was defined as ones in the following categories:

1. Managers and Senior Officials
2. Professional Occupations
3. Associate Professional and Technical Occupations
4. Administrative and Secretarial Occupations
5. Skilled Trades Occupations

Unskilled jobs were defined as ones in the following categories:

6. Personal Service Occupations
7. Sales and Customer Service Occupations
8. Process, Plant and Machine Operatives
9. Elementary Occupations

Employment status at recruitment was coded as:

1. Looking after home/family (homemaker) versus employed
2. Out of labour force versus employed
3. Retired versus employed
4. Unemployed versus employed

#### Degree Status

Degree status was coded as having a college or university degree. We did not consider professional qualifications to be equivalent to degree-level education.

### 3.1 Sensitivity Analyses

**Supplementary Table S6** contains all results for sensitivity Mendelian randomization analyses.

There was little consistent evidence in both splits of heterogeneity between SNPs in the sensitivity Mendelian randomization analyses, and little evidence of directional pleiotropy from MR-Egger regression.

There was no strong evidence for an effect of bioavailable testosterone on any outcome in any sensitivity Mendelian randomization analysis ($P > 0.01$ in all combined analyses), although effect estimates were imprecise.

Similarly, there was no strong evidence of effects of albumin, total and free testosterone on any main outcome in any sensitivity Mendelian randomization analysis ($P > 0.01$ in all combined split analyses). There was evidence of heterogeneity from MR-Egger regression between SHBG and some outcomes.

### 3.2 Secondary Analyses

**Supplementary Table S5** contains all results for main and secondary analyses. In Mendelian randomization analyses, confidence intervals were wider than multivariable-adjusted analyses, but there was little evidence for effects of any exposure on any outcome for any group. Outcomes for which there was some evidence are discussed below.

#### 3.2.1 Other Groups

##### All Women

There was no strong evidence of effects of bioavailable testosterone on any outcome for women, with or without adjusting for menopausal status.

##### Post-Menopausal Women, Not Adjusting for Oophorectomy

The results for post-menopausal women when not adjusting for oophorectomy were extremely similar to the results when adjusting for oophorectomy.

#### 3.2.2 Other Exposures

##### Total Testosterone

There was no strong evidence of effects of total testosterone on any outcome.

##### Free Testosterone

There was no strong evidence of effects of free testosterone on any outcome.

##### SHBG

There was some evidence SHBG reduced BMI in pre-menopausal women (standardised beta = -0.35, 95% CI: -0.58 to -0.11, P = 0.004).

##### Albumin

There was strong evidence from Mendelian randomization analyses that albumin reduced BMI in men (standardised beta = -0.39, 95% CI: -0.53 to -0.25, $P = 4.8 \times 10^{-8}$).

#### 3.3.3 Other Outcomes

##### Equivalisation of Household Income

We equivalised household income before tax (household income divided by the number in household, setting the maximum number in the household at 12) to explore whether household size contributed to effects on household income.

In the main Mendelian randomization analysis, using equivalised rather than raw household income did not materially change our interpretation of whether bioavailable testosterone affected household income.

### Household Income and Employment Outcomes for All Participants

In the main analysis, we restricted household income and all employment outcomes to participants of working age; in these analyses we included all participants regardless of age for these outcomes.

In the main Mendelian randomization analysis, including all participants, not just those of working age, did not materially change our interpretation of whether bioavailable testosterone affected household income or employment outcomes.
